# Unplanned readmission to hospital and its predictors in heart failure patients, Ethiopia: a retrospective cohort study

**DOI:** 10.1101/2022.11.11.22282211

**Authors:** Birhanu Ayenew, Prem Kumar, Adem Hussein

## Abstract

**Background:** The burden of heart failure increases over time and is a leading cause of unplanned readmissions worldwide. In addition, its impact has doubled in countries with limited health resources, including Ethiopia. Identifying and preventing the possible contributing factors is crucial to reduce unplanned hospital readmission and to improve clinical outcomes.

**Method:** A hospital-based retrospective cohort study design was employed from January 1, 2016, to December 30, 2020. The data was collected from 572 randomly selected medical records using data extraction checklists. Data were entered in Epi-data version 4.6 and analyzed with Stata version 17. The Kaplan-Meier and log-rank tests were used to estimate and compare the survival failure time. A Cox proportional hazard analysis was computed to identify predictors of readmission. Finally, the statistical significance level was declared at a p-value <0.05 with an adjusted odds ratio and a 95% confidence interval.

**Result:** In this study, a total of 151 (26.40%) heart failure patients were readmitted within 30 days of discharge. In the multivariate cox proportional hazards analysis being an age (>65 years) (AHR: 3.172, 95%CI:.21, 4.55), rural in residency (AHR: 2.47, 95%CI: 1.44, 4.24), Asthma/COPD (AHR: 1.62, 95%CI: 1.11, 2.35), HIV/AIDS (AHR: 1.84, 95%CI: 1.24, 2.75), Haemoglobin level 8-10.9 g/dL (AHR: 6.20, 95%CI: 3.74, 10.28), and Mean platelet volume >9.1fl (AHR: 2.08, 95%CI: 1.27, 3.40) were identified as independent predictors of unplanned hospital readmission.

**Conclusion:** The incidence of unplanned hospital readmission was relatively high among heart failure patients. Elderly patients, rural residency, comorbidity, higher mean platelet volume, and low hemoglobin level were independent predictors of readmission. Therefore, working on these factors will help to reduce the hazard of unplanned hospital readmission.

## Background

Unplanned hospital readmissions within 30-days of discharge from an index admission are widely accepted benchmarks of healthcare quality and it is a global challenge that makes healthcare quality questionable. At the same time, heart failure is the leading cause of hospital admission and readmission (1-3).

The burden of heart failure is escalating and is the greatest threat to human health and development(4). The problem of heart failure is now growing faster than our capacity to combat it, particularly in low and middle-income countries and the situation worsens, forcing African countries’ health systems to grapple with the dual burden of communicable and non-communicable diseases (5, 6). The recent international multicenter cohort study revealed that the Congestive heart failure-related mortality rate was highest in Africa (34%), particularly (42.7%) morbidity and mortality in Sub-Saharan Africa, which includes Ethiopia (7). In addition to its high morbidity and mortality in Sub-Saharan Africa (SSA), HF predominantly affects young and middle-aged people, who are more economically productive. This negatively affects multi-dimensionally the socioeconomic and the country’s development(8).

As part of the SSA country, Ethiopia shares the burden of HF. In addition, the country is complicated by a shortage of hospital beds, infrastructure, and intensive care units (9, 10). It also has the lowest health workforce density in Africa, five times below the WHO minimum threshold of 4.45 per 1000 population to meet the Sustainable Development Goal(SDG) health targets(11). These challenge the country to achieve Sustainable Development Goal3.4, which is targeted to reduce non-communicable diseases by a third in 2030. At the same time, unplanned hospital readmission upset the accessibility and quality of care(12).

The goal of treatment for heart failure patients includes reducing symptoms and disease progression, prolonging survival, and improving quality of life. However, in Ethiopia, heart failure patent came after NYHA Class III and IV with limited knowledge about their disease, poor medication adherence practice, and transitional care (13-15).

Furthermore, risky lifestyles such as smoking, a sedentary lifestyle, excessive salt intake, and alcohol consumption are most common in Ethiopia, which are predictors of unplanned hospitalizations for HF patients(16, 17). Moreover, that can be distressing and uncomfortable for the patient, his family members, and even healthcare professionals(18).

Despite this, unplanned hospital readmission is potentially avoidable through individualized care, improved clinical management, and appropriate discharge planning at index admission in collaboration with patients and family members to achieve the desired treatment goal (19, 20). However, HF patients are one the prior vulnerable than others in the first place in developed counties but not yet well characterized in sub-Saharan Africa, particularly in this study area. Therefore, Studying the incidence and predictors of unplanned hospital readmission among heart failure patients is vital for health policymakers to support quality improvement efforts by developing preventive guidelines and treatment protocols to reduce the incidence of readmission and related consequences.

## Methods and materials

### Study design, period, and area

A hospital-based retrospective follow-up study was conducted from January 1, 2016, to December 31, 2020. This study includes all general hospitals of South Wollo Zone, namely Boru-Meda General Hospital, Akesta General Hospital, and Mekane-Selam General Hospital, which are functional during the follow-up period.

### Inclusion Criteria

All heart failure patients hospitalized from January 1, 2016, to December 31, 2020, and whose age is ≥ 18 years old were included.

### Exclusion Criteria

Heart failure patients didn’t have baseline data, and patients discharged with death were excluded from this research to avoid estimation bias.

### Sample size determination and sampling procedure

The maximum sample size of this study, obtained from predictors for Cox proportional hazard (PH) regression, calculated using the software STATA version 17.0 assuming a 95% confidence interval, significance level (5%), power 80%, withdrawal effect (15%) and adjusted hazard ratio on being a commodity of diabetes mellitus taken from (11) giving a total sample size of 626. You can see details the sampling procedure in the figure below (figure 1).

**Figure 1.**
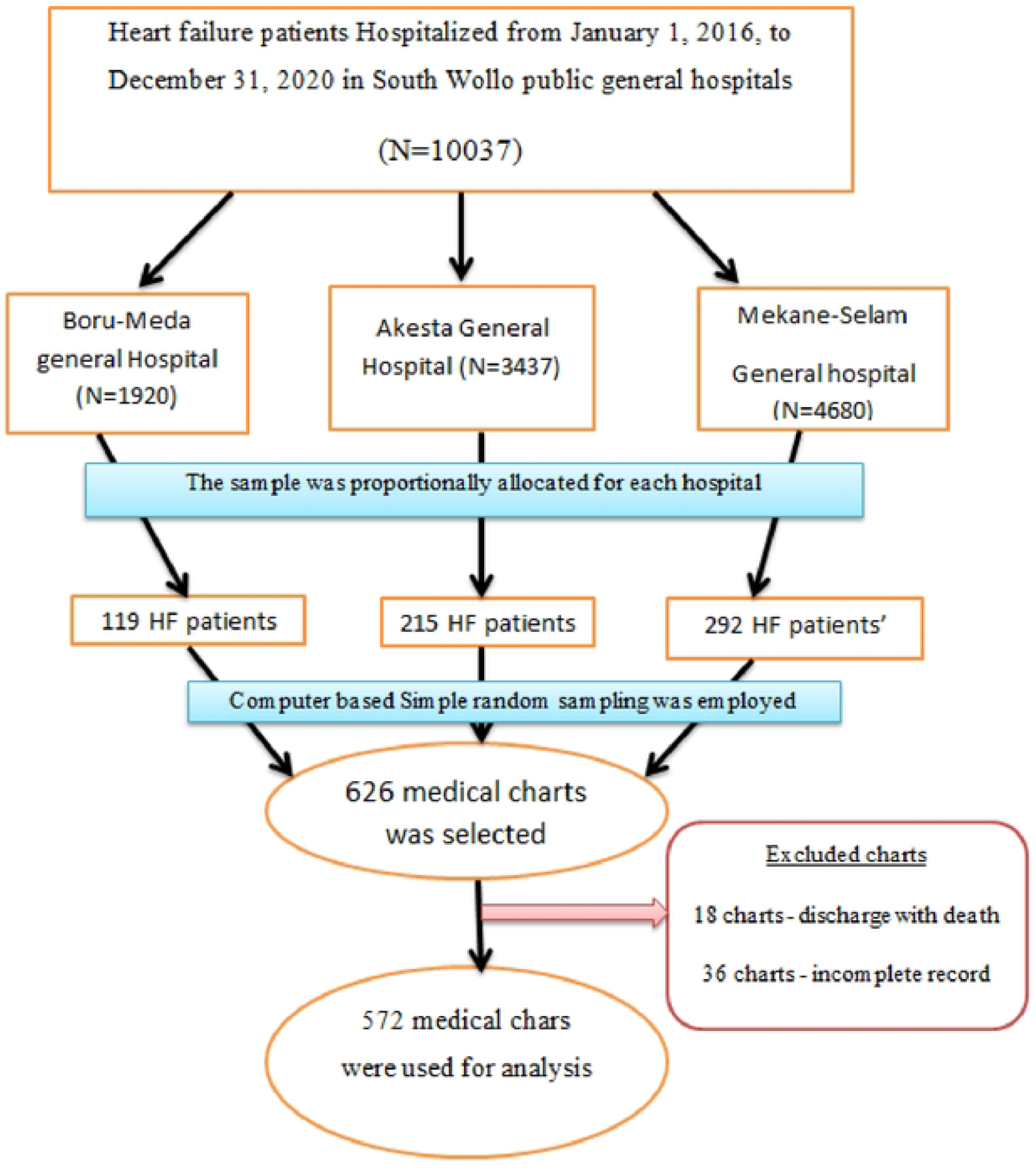
Schematic representation of sample selection procedure of study participants in heart failure patients with at selected South Wollo general Hospitals Ethiopia, 2022.

### Data collection tools, procedures, and quality control

The data was collected using data extraction tools adapted by reviewing different articles (21, 22). The list of patients hospitalized with index admission of HF was retrieved from the HMIS registration using the medical registration numbers. 3 BSc nurses were involved in data collection, and supervision was done. The data extraction tool was checked for its completeness and existence of relevant variables through a preliminary chart review of 10% of the sample size at Kemisse General Hospital. And the data collectors were trained on the data collection process before the actual data collection period.

### Operational definition

#### Event

The occurrence of readmission within 30 days after hospital discharge from an index admission.

#### Follow-up time

From the time of discharge until an event occurred

#### Censored

Patients who did not readmit within 30 days during the follow-up period

#### Survival status

The status of heart failure patients at the end of the follow-up period (readmission or censored)

#### Time to readmission

The Time interval from discharge from the hospital till readmission happens

#### Incomplete patient data on chart

refer to charts with no discharge date, date of readmission, and laboratory results of complete blood count(CBC) at discharge during the index admission.

##### Data processing and analysis

After data collection, its completeness was checked before data entry, and then Epi-data manager version 4.6 was used for data entry and explored to Stata version 17 for analysis. The Kaplan-Meir curves were used to estimate the survival status, and the log-rank test was used to compare the association between the outcome and predictor variables.

The bi-variable analysis was computed between the dependent and each independent variable, and variables with p-value≤ 0.2 at the bi-variable were included in the multivariable analysis to identify independent predictors of readmission. The Cox proportional hazard model’s fitness was assessed using Schoenfield residual and global tests, and the p-value was insignificant. A multivariable Cox-hazard proportional analysis was computed to estimate the effect size of predictor variables on the outcome variables with a 95% confidence interval. Then, a p-value ≤ 0.05 was considered statistically significant.

### Ethical Considerations

The Ethical Clearance Letter was obtained from Wollo University’s Ethical Review Committee. It has been given an IRB number of CMHS/03/14. Then officials at different levels in the hospitals were communicated through letters, and permission was assured. The data was not disclosed to any person other than the principal investigator. The entire study was conducted as per the declaration of Helsinki’s ethical principles for medical research.

## Results

Among 572 study participants, 302(52.8%) were male, and 370(64.7%) were rural in residency. Regarding age distribution, the median age of the study participants was 45 years, and the mean age was 45.8± 14.1 SD years. (See details from table 1 below).

**Table 1.** Socio-demographic characteristics of adult heart failure patients at selected South Wollo General Hospitals in 2022

| Variables | Category | Status |  | Total (%) |
| --- | --- | --- | --- | --- |
|  |  | Readmitted (%)<br>(n=151) | Censored (%)<br>(n=421) |  |
| Age | <65 | 103(68.21) | 388(92.16) | 491(85.84) |
|  | ≥65 | 48(31.79) | 33(7.84) | 81(14.16) |
| Sex | Male | 81(53.64) | 221(52.49) | 302(52.80) |
|  | Female | 70(46.36) | 200(47.51) | 270(47.20) |
| Residency | Urban | 37(24.50) | 165(39.19) | 202(35.31) |
|  | Rural | 114(75.50) | 256(60.81) | 370(64.69) |

### Baseline laboratory marker of the study participants on discharge

Concerning baseline laboratory markers of the study participants, from study participants, a Platelet count greater than 150 cells/ L 136 (90.07%) was readmitted, and White blood cell count (103 /µL) from ≥1.3 to ≤4 is 122(80.79%) was readmitted. Participants with a mean platelet volume (fl) greater than 9.1 were readmitted 116(76.82%). You can see details in the table below (Table 2).

**Table 2.** Baseline laboratory marker of the study participants during index admission in heart failure patients with at selected South Wollo general Hospitals; Ethiopia, 2022.

| Variables | Category | Status |  | Total (%)<br>(n=572) |
| --- | --- | --- | --- | --- |
|  |  | Readmitted (%)<br>(n=151) | Censored (%)<br>(n=421) |  |
| White blood cell<br>(10<sup>3</sup> /μL) | <1.3 | 17(11.26) | 51(12.11) | 68(11.89) |
|  | ≥1.3 - ≤4 | 122(80.79) | 329(78.15) | 451(78.85) |
|  | >4 | 12(7.95) | 41(9.74) | 53(9.27) |
| Platelet count<br>(cells/ L) | ≤150 | 15(9.93) | 162(38.48) | 177(30.94) |
|  | >150 | 136 (90.07) | 259(61.52) | 395(69.06) |
| Mean platelet<br>volume (fl) | <9.1 | 35(23.18) | 292(69.36) | 327(57.17) |
|  | ≥9.1 | 116(76.82) | 129(30.64) | 245(42.83) |
|  | ≥13 | 21(13.91) | 299 (71.02) | 320(55.94) |
| Hgb(g/dL) | 11-12.9 | 36(23.84) | 59 (14.01) | 95(16.61) |
|  | 8-10.9 | 94(62.25) | 63 (14.95) | 157(27.45) |

### Preexisting comorbidity status of study participants

Regarding preexisting comorbidities status, from study participants, hypertension is the leading in 106(18.53%), followed by COPD/Asthma 95(16.61%). You can see details in the table below (Table 3).

**Table 3:**
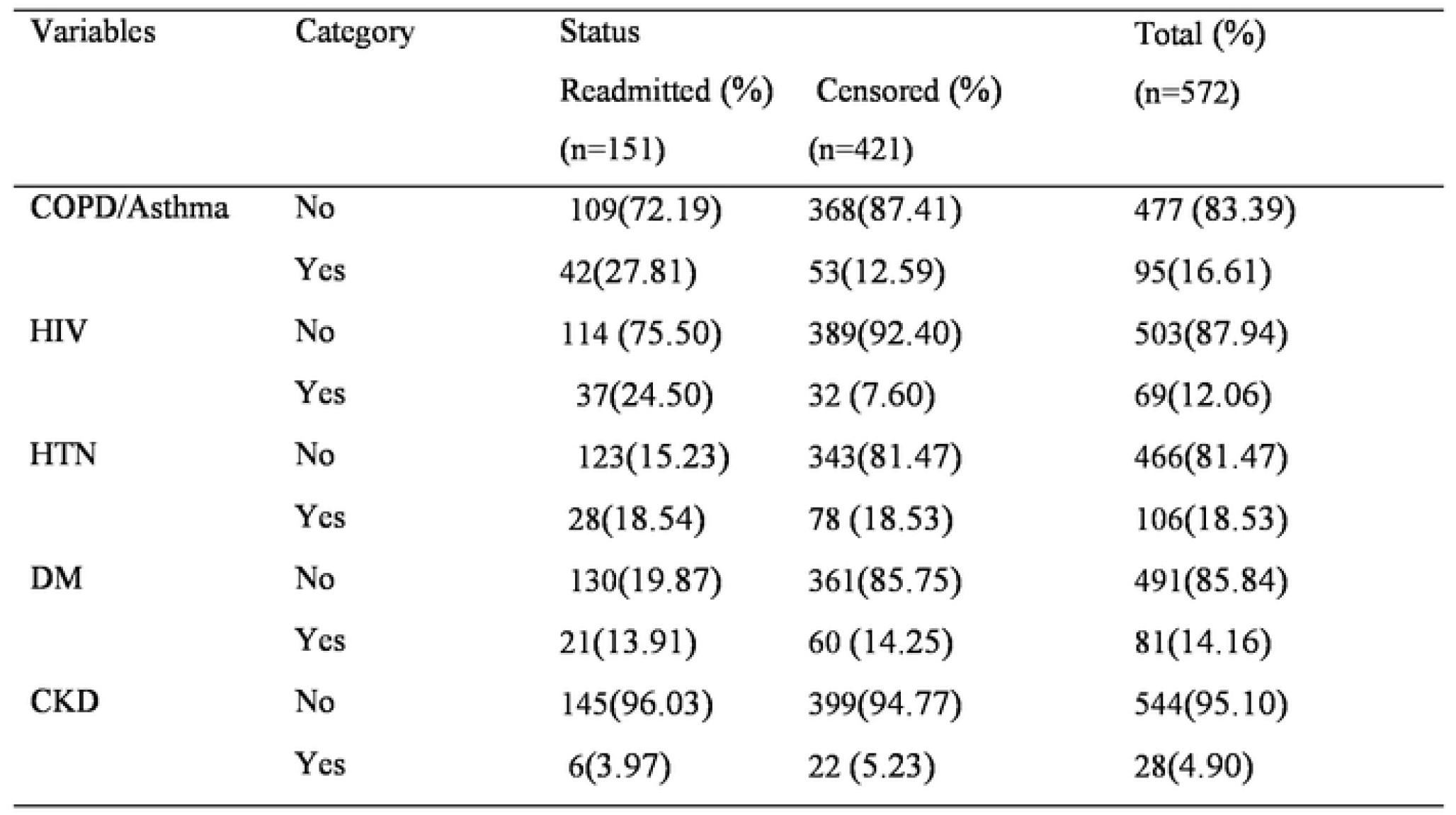
Pre-existing co-morbidity status of study participants in heart failure patients with at selected South Wollo general Hospitals Ethiopia, 2022.

### The incidence of unplanned hospital readmission in heart failure patients

In this study, 572 adult heart failure patients were followed retrospectively. The median time of readmission was 16 days (95% CI: 14, 17), with a minimum of 3 days and a maximum of 30 days of follow-up time. In this study, 421 patients were censored, and 151 were readmitted within 30 days of discharge, resulting in a cumulative incidence of readmission of 26.40% (95% CI: (23.0, 30.2) during the follow-up period. The total follow-up time was 15,093 person-day, with an incidence rate of 10.01 readmission per 1000 person-day observations (95% CI: 8.79, 13.38). You can see from (figure 2)

**Figure:2.**
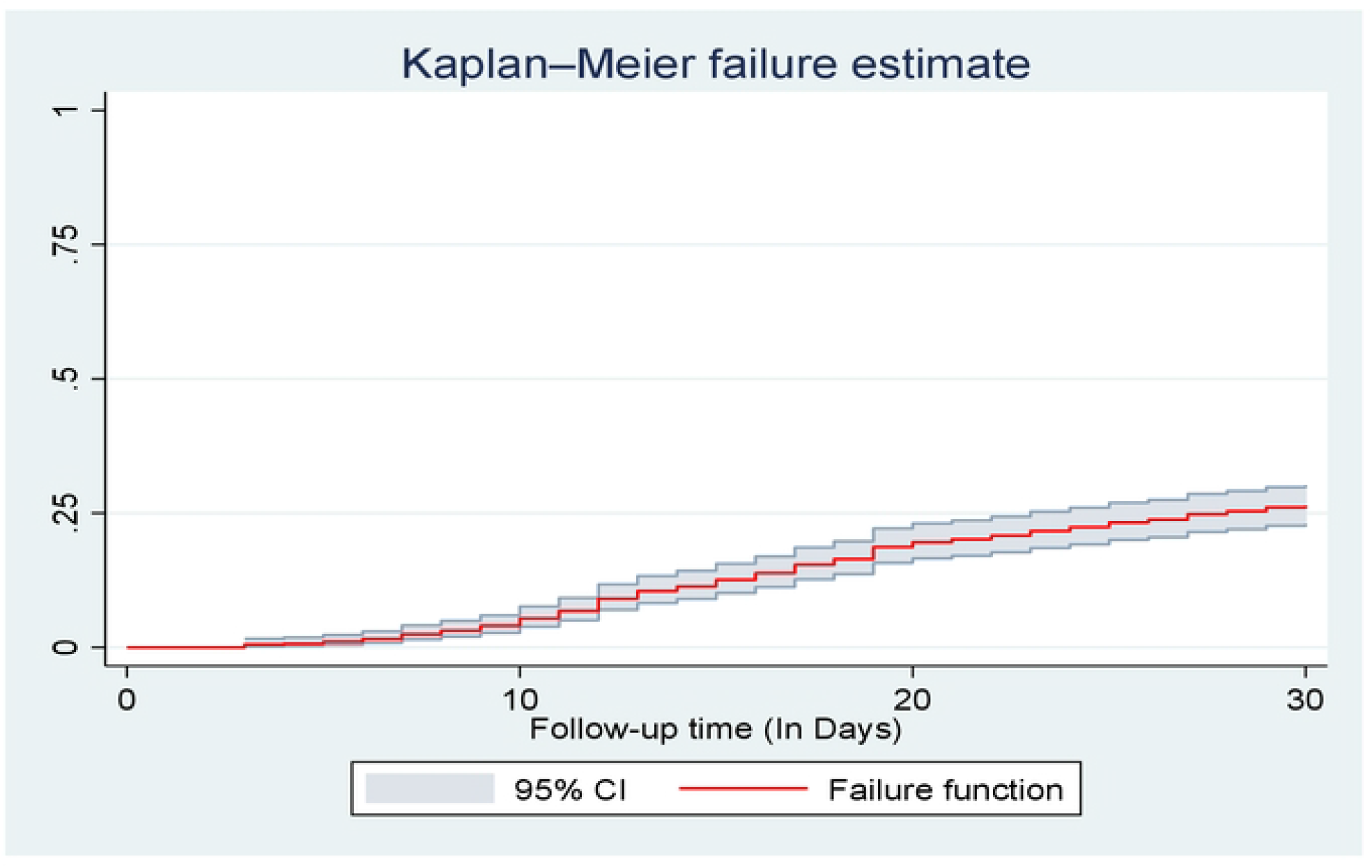
the overall Kaplan-Meier analysis of the hazard of unplanned readmission in heart failure patients at selected South Wollo general Hospitals; Ethiopia, 2022.

**Figure: 3.**
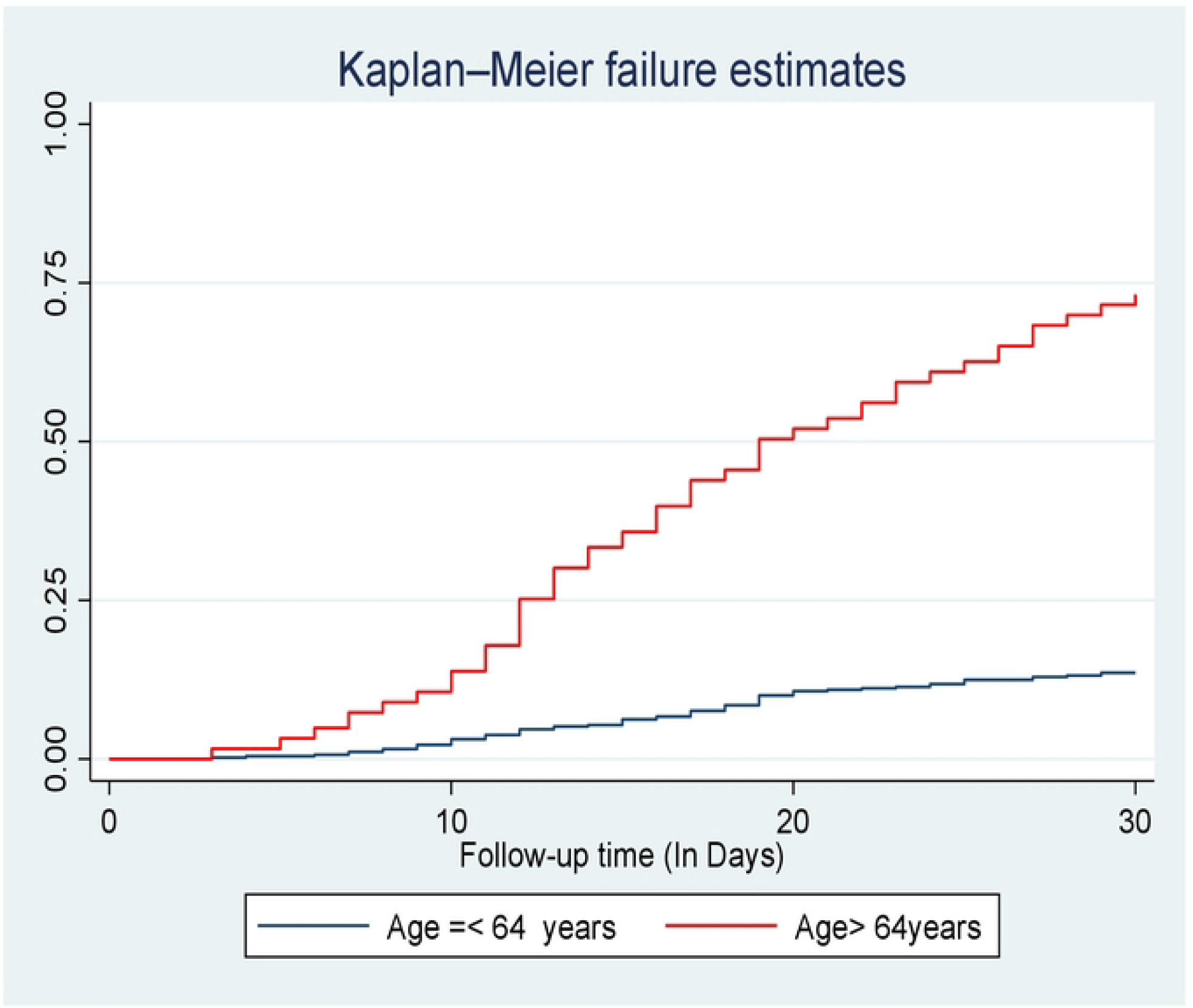
kaplan- meier hazard estimation of unplanned readmission in heart failure patients with categories of age at selected South Wollo general Hospitals; Ethiopia, 2022.

**Figure:1.**
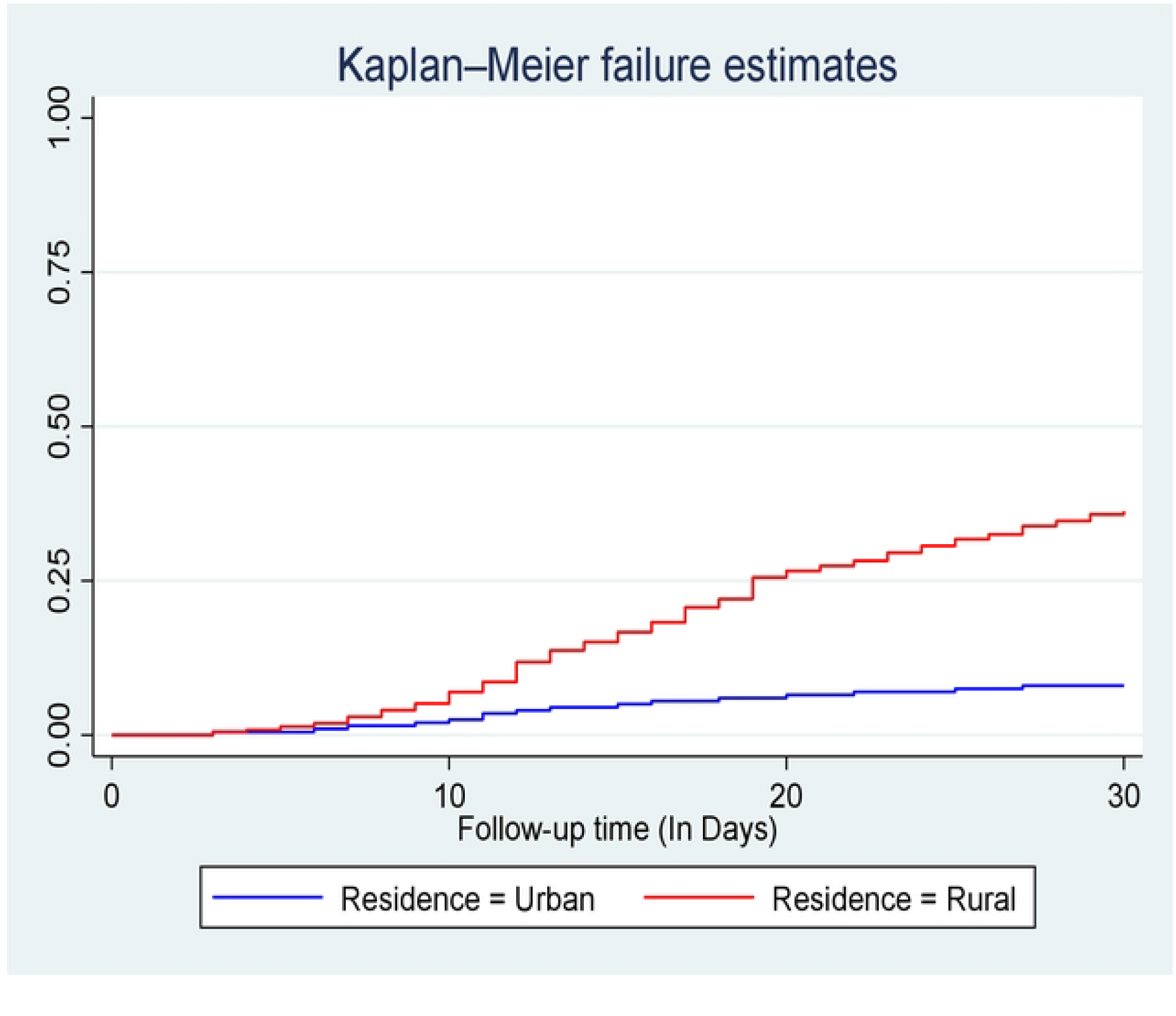
kaplan- meier hazard estimation of unplanned readmission in heart failure patients with categories of residence at selected South Wollo general Hospitals; Ethiopia, 2022.

**Figure:5.**
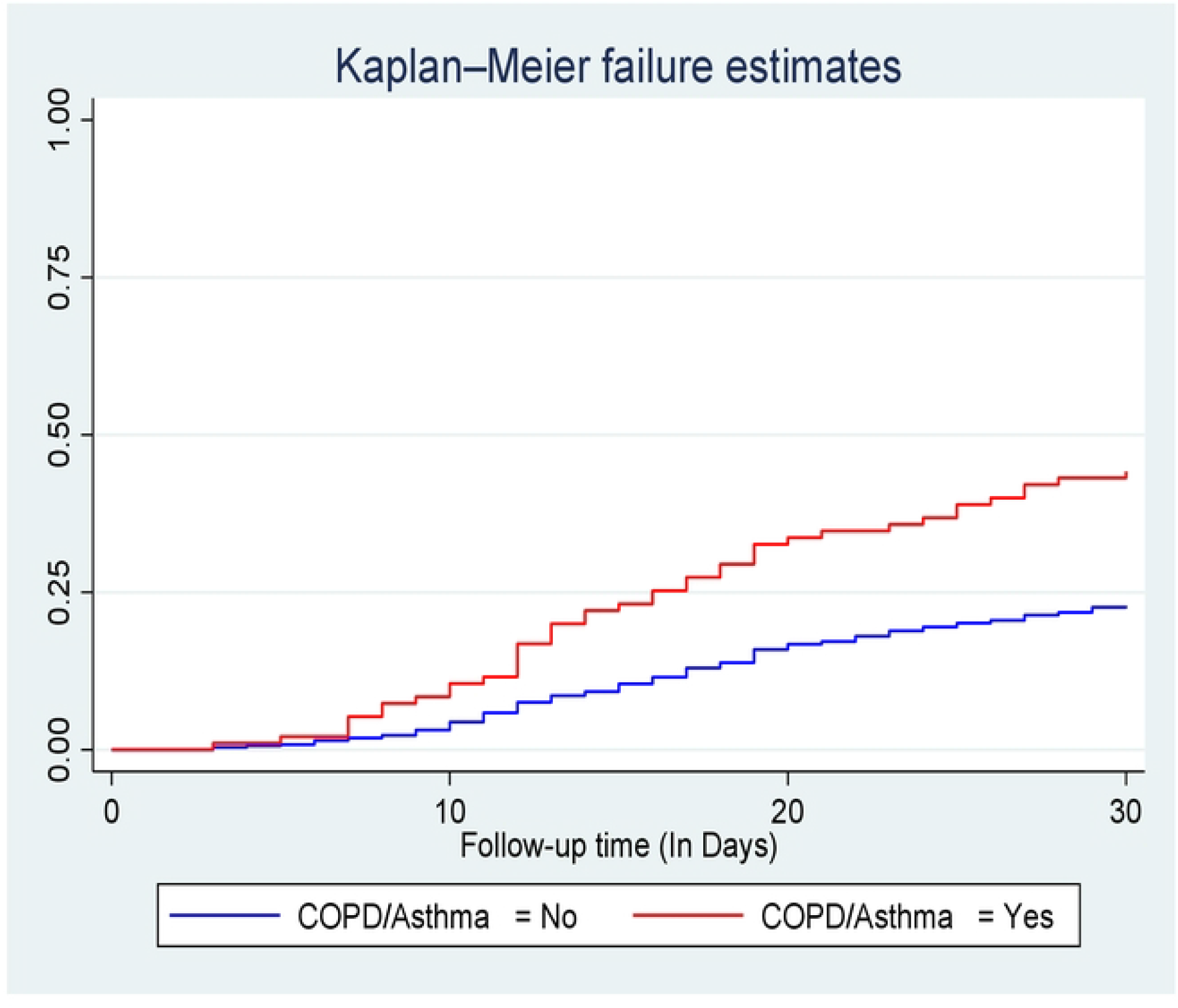
kaplan- meier hazard estimation of unplanned readmission in heart failure patients with categories of COPD/Asthma at selected South Wollo general Hospitals; Ethiopia, 2022.

**Figure:6.**
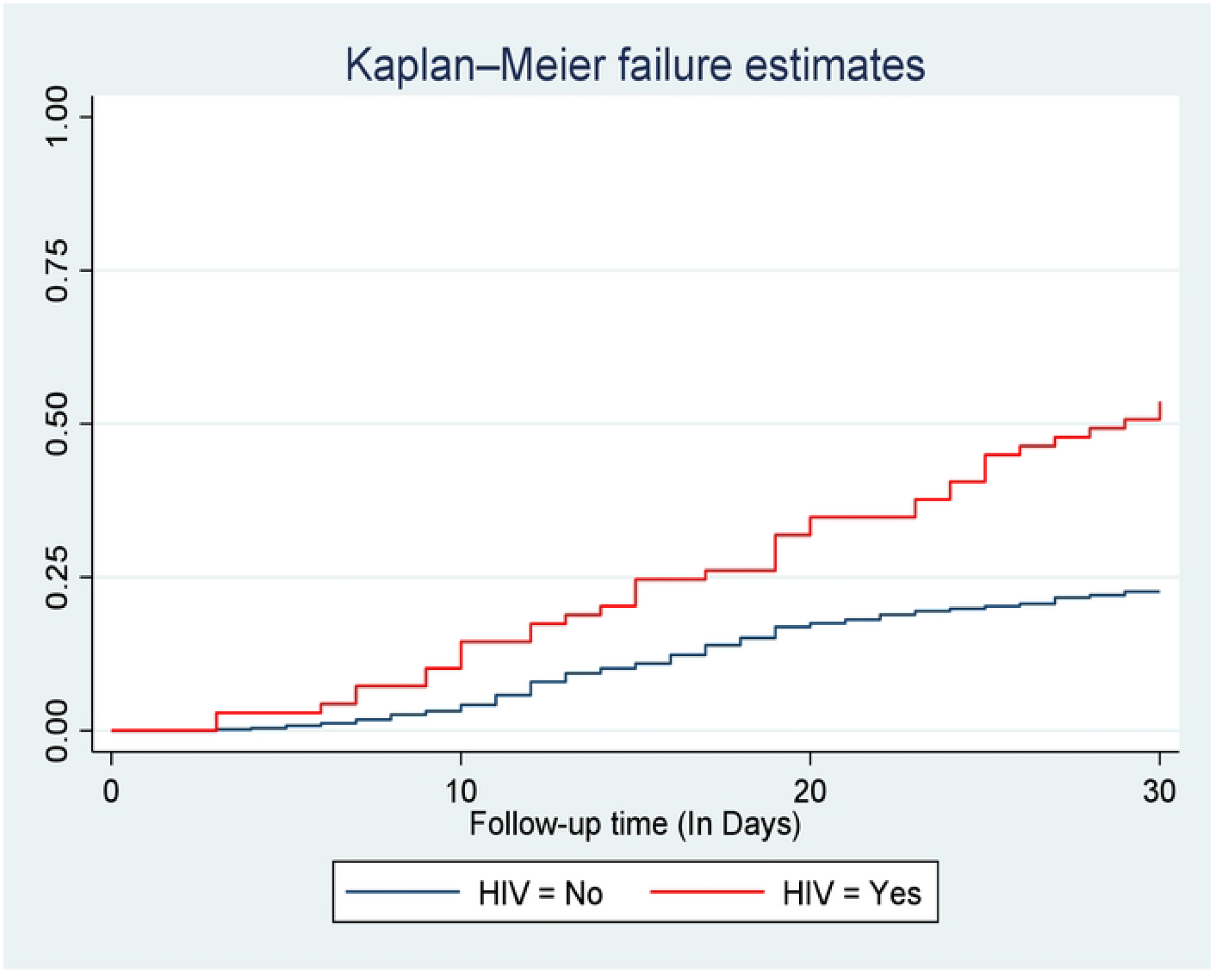
kaplan- meier hazard estimation of unplanned readmission in heart failure patients with categories of HIV/AIDS at selected South Wollo general Hospitals; Ethiopia, 2022.

**Figure:7.**
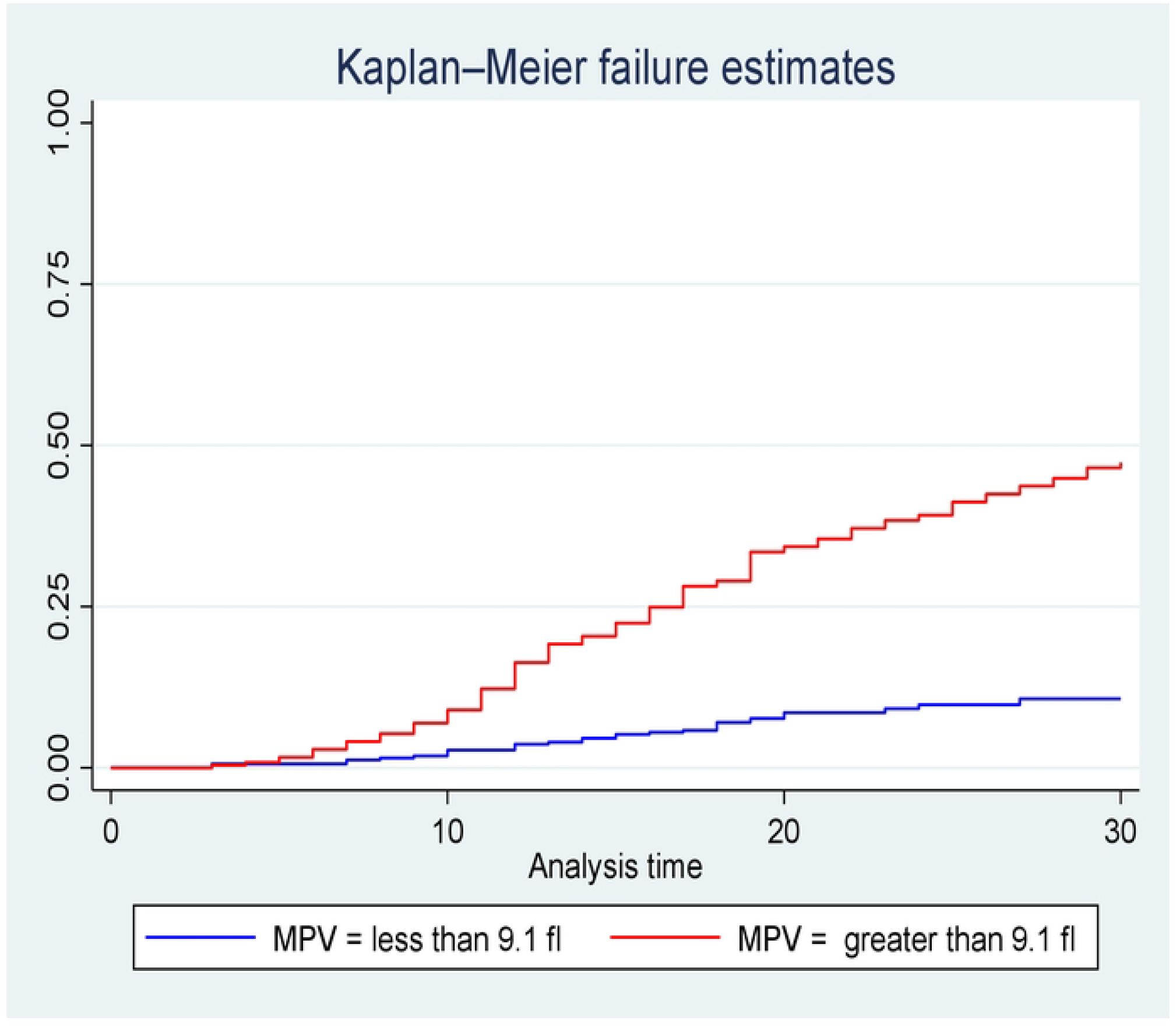
kaplan- meier hazard estimation of unplanned readmission in heart failure patients with categories of mean platelet volume **(MPV)** at selected South Wollo general Hospitals; Ethiopia, 2022.

**Figure:8.**
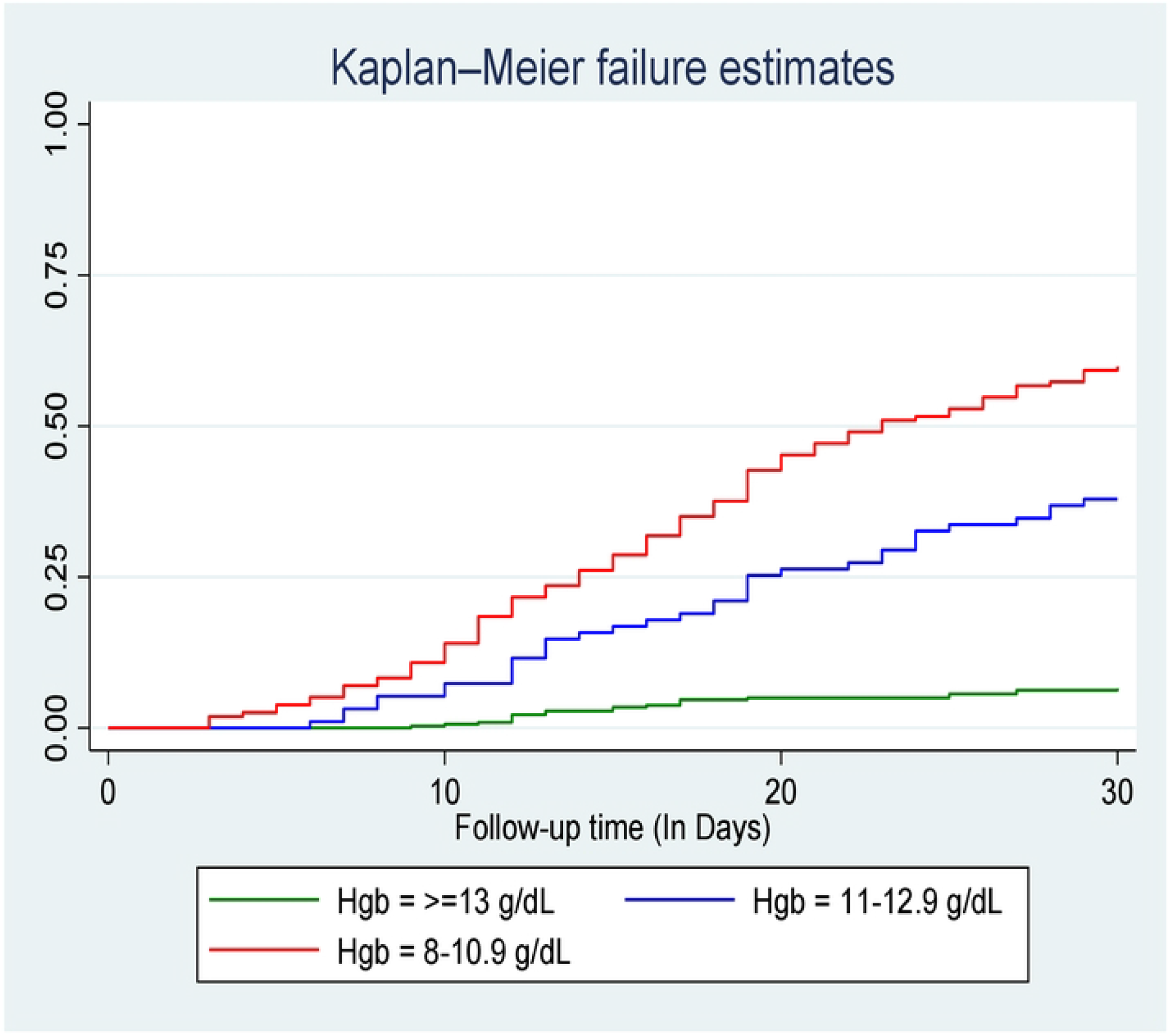
kaplan- meier hazard estimation of unplanned readmission in heart failure patients with categories of haemoglobin (Hgb)at selected South Wolle general Hospitals; Ethiopia, 2022.

### Kaplan- Meier hazard estimation of unplanned readmission in heart failure patients

The overall Kaplan- Meier estimate showed that the hazard of unplanned hospital readmission of heart failure patients is low during the first three days following discharge after index admission. However, which relatively increases the hazard of unplanned readmission as follow-up time increases. The overall median time of unplanned readmission among heart failure patients was 16 days (95% CI: 14, 17), and the meantime of the study participant was 26.38 days (95% CI: 23.82, 30.63). The hazard of unplanned readmission of heart failure patients at the start of follow-up was 100%. The hazard of unplanned readmission at three days, five days, and ten days were 0.52%, 1.05%, and 5.42%, respectively.

During follow-up time following discharge from the hospital during index admission to 30 days, the hazard curve tends to rise rapidly, implying unplanned reemitted in heart failure patients within this period.

### Predictors of unplanned hospital readmission in heart failure patients

Cox proportional hazard regression model was computed to identify the relationship between the hazard of readmission and independent variables. In the Bi-variable Cox-Proportional Hazards regression model; sex, age, HIV/AIDS, COPD/Asthma, Haemoglobin level (Hgb), mean platelet volume (MPV), and platelet level were found to be p-value of less than 0.25 with unplanned hospital readmission. Those variables having a p-value of <0.25 in the Bi-variable analysis were fitted in multivariable analysis.

In the multivariable cox-proportional hazards model, age, HIV/AIDS, COPD/Asthma, Haemoglobin level (Hgb), and Mean Platelet Volume (MPV) were Significant predictors of unplanned hospital readmission with a P-value of <0.05.

The multivariable Cox-Proportional Hazards analysis revealed that patients with heart failure, who were age ≥65 years had more than three-fold hazard of unplanned readmission (AHR: 3.172, 95%CI: 21, 4.55) than those heart failure patients in the age group between 18 and 64 years. The hazard of unplanned readmission among heart failure patients was 2.47 times (AHR: 2.47, 95%CI: 1.44, 4.24), higher for rural residents than for urban.

Furthermore, Patients with baseline Haemoglobin level 11-12.9 g/dL and Haemoglobin level 8=10.9 g/dL had more than three times and six times at high hazard of unplanned readmission (AHR 3.67, 95%CI: 2.09, 6.44), and (AHR: 6.20, 95%CI: 3.74, 10.28)than those with baseline hemoglobin level >13 g/dL during index admission respectively. You can see details in the table below (Table 4).

**Table 4.**
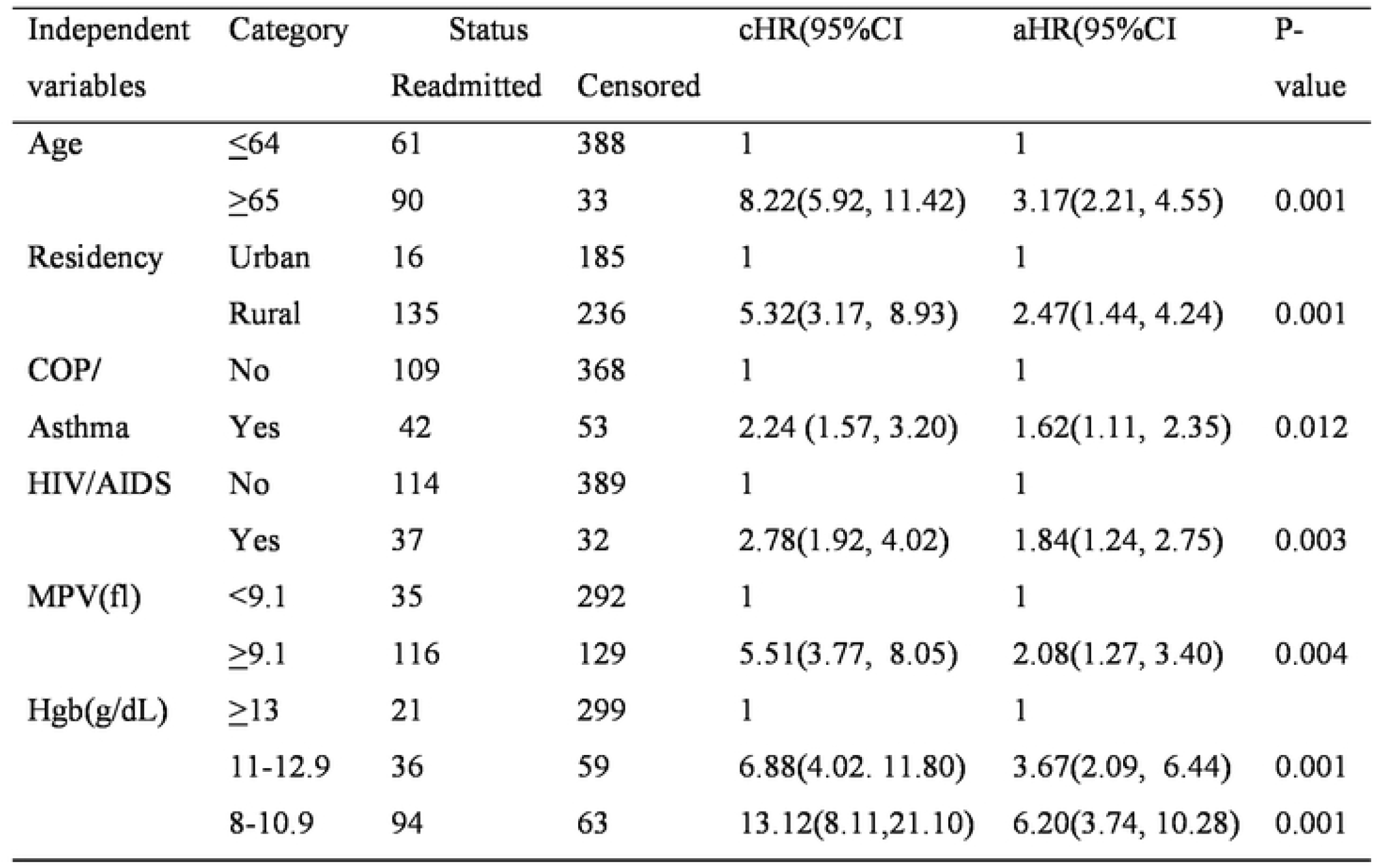
Cox proportional hazard regression model of the study of in heart failure patients with at selected South Wallo general Hospitals Ethiopia, 2022

## Discussion

This study assessed incidence and predictors of 30-day unplanned hospital readmission in heart failure patients at selected South Wollo General Hospitals; Ethiopia, 2022. This study found that the cumulative overall incidence of unplanned hospital readmissions during the follow-up period was 26.40%; 95% CI: (23.0, 30.2). This result is quite close to the study conducted in the USA, which is 29.7%(23), and unplanned readmission across Africa, which was 24.3% (24).

On the other hand, the cumulative overall incidence of unplanned readmissions is higher in this study than study conducted in Iran, which is 6.6%(25), 20.6% in Nigeria, 13.3 % Mozambique, 16.0% South Africa, 22.3% in Australia and New Zealand and 6.56% in Japanese(1, 24, 26). This difference might be due to demographic characteristics. Furthermore, the discrepancy might be due to the follow-up period difference. A study conducted in Mozambique, Nigeria, South Africa, and Japan was a 1-year cohort study; in addition, the study conducted in Iran was a 5-month study. Both had short follow-up times and may have been influenced by different factors, but the current study contains 5-year data.

Also, the finding of this study is lower than the study conducted in Sudan, where is 37.3% of heart failure patients are readmitted(24). This difference might be due to operational definition deference. This study incorporates readmission if patients are readmitted within 30 days, but the Sudan study counts as readmission if readmitted within one year.

This study revealed that being older age >65 increases the hazard of unplanned hospital readmission as compared to adults in the age group of 18-64. This finding is supported by a study conducted in Japan(27). The possible reason might be that older patients are particularly vulnerable to physiological deterioration, comorbidity, and psychological stress(28). Additionally, the risk of errors in medication intake and a sedentary lifestyle increased in elderly patients (29). Indeed, we found that older patients were more likely to be readmitted with non-heart failure-related conditions since the operational definition of this study incorporates all causes of readmission. And this implies that older patients are highly vulnerable to readmission and need particular attention.

This study found that being rural residents was an independent predictor of unplanned hospital readmission. This finding is supported by a study conducted in Canada(30) and Chile(31), which concluded that rural heart failure patients are at risk for frequent hospitalizations due to poor self-care and poor knowledge of heart failure. The possible reason might be that rural heart failure patients are slower to adopt healthy behaviors and have lower health literacy levels than residents of urban communities(32). Interventions aimed at expanding disease-related knowledge in patients with heart failure can positively impact re-hospitalization and quality of life.

In this study, those patients with heart failure during index admission with preexisting COPD/Asthma increased the hazard of unplanned readmission compared to patients without COPD /Asthma. This finding is supported by the study conducted in the US(33), Sweden(22), and Massachusetts General Hospital(34), which reported that having respiratory disease increases the risk of 30-day readmission. And These findings were supported by (35)and (36) conclude that respiratory infections are a known risk factor for heart failure, and low oxygen levels due to COPD/Asthma cause blood pressure to rise in the arteries of the lungs and cause a reduction in the ability to pump blood, putting extra strain on the heart and worsening symptoms and might be cause unplanned hospital readmission. Furthermore, this study incorporates all causes of readmission and might be reemitted non-HF-related disease. And this implies that HF patients with COPD/Asthma are highly vulnerable to unplanned hospital readmission and need particular attention.

In this study, heart failure patients living with HIV/AIDS had a higher risk of unplanned hospital readmission than patients without HIV/AIDS. This finding is supported by other studies in the United States (37), Massachusetts (38), and New York City(39) were reported that preexisting HIV/AIDS HIV infection increases the risk of unplanned readmission in patients with heart failure.

This might be due to the fact that living with HIV/AIDS is directly associated with low immune function, thereby increasing the risk of both bacterial and viral infection and, consequently, poor outcomes(40). This finding highlights the importance of timely monitoring and greater conservative management for those individuals with these comorbidities. Furthermore, optimal antiretroviral therapy is essential for patients with HIV who develop HF. Effective antiretroviral treatment that results in immunological rebound and viral suppression may partially protect people living with HIV who also have heart failure from the adverse events of heart failure.

This study found that those patients who had lower hemoglobin levels at discharge during index admission increased the risk of unplanned readmission to the hospital compared to a normal hemoglobin level. This study is consistent with the study conducted by the European Society of Cardiology (41), the United Arab Emirates (42), India(43), and the meta-analysis of 26 studies in patients with the comorbidity of heart failure and anemia(44).

Anemia is a known cause of deterioration in heart function, as it causes both cardiac stresses from tachycardia and increases stroke volume, which can lead to reduced renal blood flow and fluid retention, which puts additional pressure on the heart. Prolonged anemia from any cause can cause left ventricular hypertrophy (LVH), which can lead to cardiac cell death through apoptosis and worsen CHF (45). This implies that heart failure patients with lower hemoglobin levels during index admission need special attention to non-pharmacological treatment in addition to pharmacological intervention, particularly hemoglobin-enhancing nutrients. In addition, patients with lower hemoglobin levels need close follow-up by shortening the follow-up time.

The present study found that an increase in mean platelet volume (MPV) of greater than 9.1 fl during index admission was an independent predictor of an increase in the risk of unplanned hospital readmission in patients with heart failure. This study is consistent with the study conducted in Turkey(46), Germany(47), and Romania(48). This may be explained as MPV being one of the potential biomarkers of platelet activity. An increase in MPV is an indicator of larger and more reactive platelet, which aggregates, represents a major risk factor for atherothrombosis and increased risk of infarction(49). This implies that MPV is a simple marker of platelet functional status and may represent a risk factor for adverse vascular events. In addition, MPV is an easy and routine blood test to perform; therefore, healthcare professionals should evaluate it and use it as a marker of prognostic factors.

### Limitations of the study

The data were derived retrospectively from medical reports of the patients and some important variables like personal behavior such as smoking, alcohol use, and chat chewing, which were strong predictors of unplanned hospital readmission in other studies but was not or inadequately recorded and not included in the analysis.

Patients discharged from one hospital during index admission may admit within 30 days from other hospitals but not recorded as readmission due to the retrospective nature of the study and hospitals not being linked. In these circumstances, the readmission rate might be underestimated.

## Conclusion

The incidence of unplanned hospital readmission among heart failure patients is relatively high. The hazard of readmission was higher among rural residents, elderly patients aged≥65 years, patients living with HIV/AIDS, COPD/Asthma, increased mean platelet volume (MPV), and a decrease in hemoglobin level during index admission were independent predictors of unplanned hospital readmission among patients with heart failure.

## Data Availability

All relevant data are within the manuscript and its Supporting Information files.

## Abbreviations

HF: Heart Failure
HMIS: Health Management Information System
MPV: mean platelet volume
SDG: Sustainable Development Goal
SSA: Sub-Saharan Africa

## Acknowledgment

First of all, we would like to thank Wollo University College of medicine and health science, school of nursing, and midwifery, department of Adult health nursing for the chance it for giving me this chance to conduct this research.

Finally, we thanks to Boru-Meda general hospital, Akesta general hospital, and MekaneSelam general hospital for quality unit control, HMIS coordinators, card room officers, and data collectors for their cooperation.

